# Learning Phenotypic Associations for Parkinson’s Disease with Longitudinal Clinical Records

**DOI:** 10.1101/2020.03.15.20036657

**Authors:** Weishen Pan, Chang Su, Kun Chen, Claire Henchcliffe, Fei Wang

**Affiliations:** State Key Laboratory of Intelligent Technologies and Systems, Beijing National Research Center for Information Science and Technology (BNRist), Institute for Artificial Intelligence (THUAI), Tsinghua University, Beijing, China; Department of Healthcare Policy and Research, Weill Cornell Medical College, Cornell University, New York, NY, USA; Department of Statistics, University of Connecticut, Storrs, CT, USA; Center for Population Health, University of Connecticut Health Center, Farmington, CT, USA; Department of Neurology, Weill Cornell Medical College, New York, NY, USA

## Abstract

**Background:** Parkinson’s disease (PD) is associated with multiple clinical manifestations including motor and non-motor symptoms, and understanding of its etiologies has been informed by a growing number of genetic mutations, and various fluid-based and brain imaging biomarkers. However, the precise mechanisms by which these phenotypic features interact remain elusive. Therefore, we aimed to generate the phenotypic association graph of multiple heterogeneous features within PD to reveal pathological pathways of the complex disease.

**Methods:** A data-driven approach was introduced to generate the phenotypic association graphs using data from the Parkinson’s Progression Markers Initiative (PPMI) and Fox Investigation for New Discovery of Biomarkers (BioFIND) studies. We grouped features based on the structure of the learned graphs in both cohorts, and investigated their dynamic patterns in the longitudinal PPMI cohort.

**Findings:** 424 patients with PD from the PPMI study and 126 patients with PD from the BioFIND study were available for analysis. For PPMI, the phenotypic association graphs were generated at different time points of the disease, including baseline (without any PD treatments), and 1-, 2-, 3-, 4-, and 5-year follow-up time points. Based on topological structure of the learned graph, clinical features were classified into homogeneous groups, that were densely intra-connected while sparsely inter-connected. Importantly, we observed both stable and longitudinally changing relations in the graphs generated, likely reflecting the dynamic pathologies of PD. By cross-cohort comparison, we observed very similar structure for graphs constructed from BioFIND (in which patients have a much longer duration of PD at enrollment than PPMI) and later-period (4- and 5-year follow-up) data from PPMI. This consistency demonstrates the effectiveness of our method.

**Interpretation:** We analyzed the heterogeneous features of PD by generating the phenotypic association graphs. By analyzing the structural relationships among the features over time, our findings could improve the understanding of the pathologies of PD.

**Funding:** Michael J Fox Foundation for Parkinson’s Research.

## Introduction

Multiple identifiable and quantifiable phenotypic features are associated with Parkinson’s disease (PD), including biomarkers, motor manifestations (such as bradykinesia, muscle rigidity, tremor, and postural instability) and non-motor manifestations (such as depression, cognitive decline, fatigue, dysautonomia, and other signs and symptoms). Although how they are linked is as for the most part quite poorly understood, and their heterogeneity between individual patients diagnosed with PD leads to further complexity, these features are related to the spatially, temporally, and molecularly complex pathologies of PD.

In order to uncover the underlying mechanisms of PD, there is an urgent need for identifying relationships among its heterogeneous phenotypic features. Typically, most of the existing cohort studies focus on a single specific feature and its relationships to other factors,^1–4^ relying on statistical testing, univariate or multivariate regression. However, these approaches require hypotheses of the independent and dependent features to be tested, and hence may not be appropriate to detect the complex correlations among the heterogeneous features of PD (see Figure 1).

**Figure 1:**
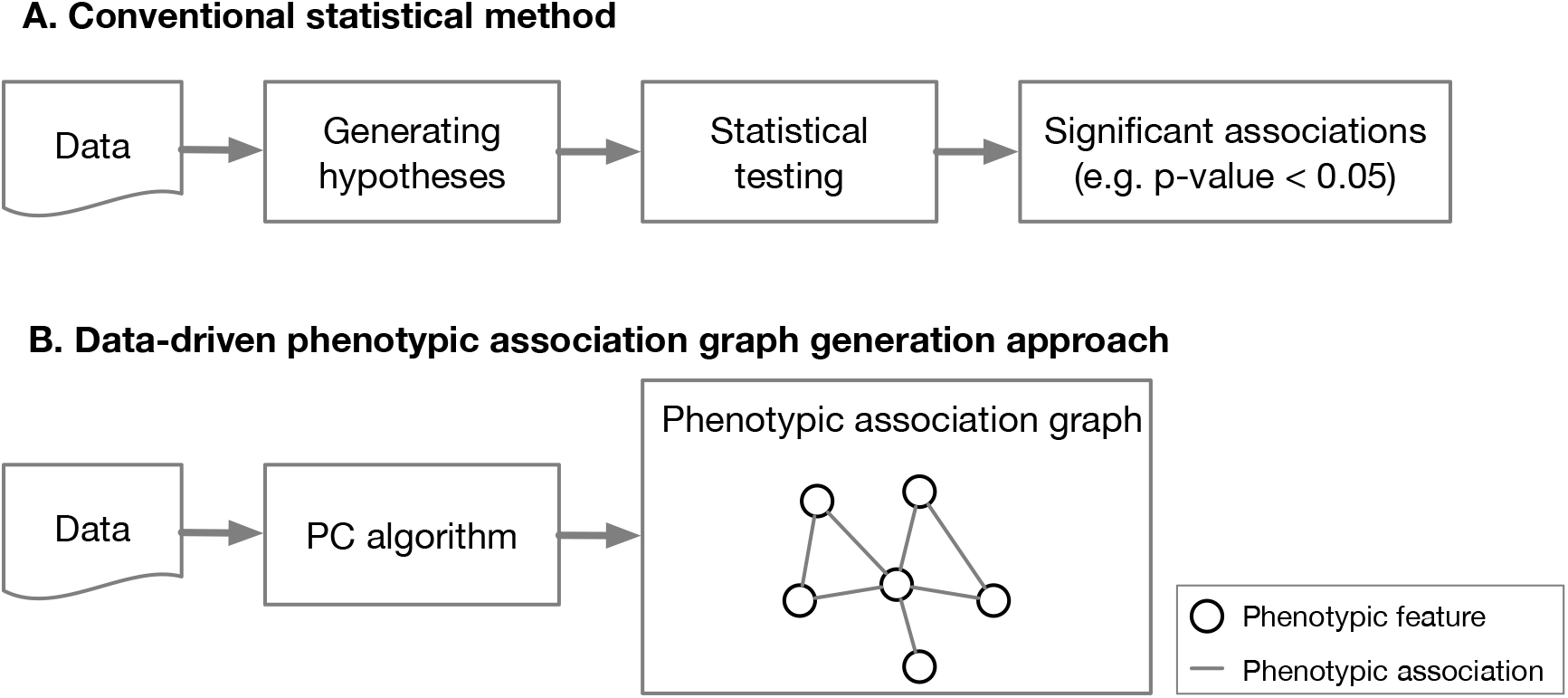
An illustration of conventional phenotypic association identification methods (A) and data-driven phenotypic association graph generation approach. Given the data, the conventional methods typically first generate some hypotheses, i.e., candidate feature associations of interest, then apply statistical testing approaches to calculate the p-values. In contrast, the data-driven approach, without any hypothesis, takes all features as input, and comprehensively generates the phenotypic association graph, of which nodes are features studied and undirected edges are identified associations among the features.

To address the issue above in PD, we propose to use a data-driven approach, PC algorithm, which takes input as all phenotypic features and produce an undirected graph, i.e., the phenotypic association graph, revealing the subtle relationships among the features (see Figure 1). For cross-cohort validation, we calculated the phenotypic association graphs within two PD cohorts. Moreover, by investigating the phenotypic association graphs generated at different periods of PD, we analyzed how the relations evolve as PD progresses. Finally, based on the obtained phenotypic association graphs, we sought to discuss the pathologies of PD.

## Method

### Study population

We used publicly available data from two cohorts comprising individuals with PD and healthy control (HC) subjects, the Parkinson’s Progression Markers Initiative (PPMI) and the Fox Investigation for New Discovery of Biomarkers (BioFIND).^5,6^

The PPMI study is a prospective longitudinal study of de novo PD patients who were untreated at baseline, and were enrolled at 33 sites internationally.^5^ Data downloaded from https://www.ppmi-info.org/data on May 03, 2018, were used for this analysis. At that time, enrollment was complete. A total of 424 individuals with PD, whose baseline data were available, were included for analysis. Follow-up assessments included complete neuropsychological test data, at 1-year, 2-years, 3-years, 4-years, and 5-years after baseline.

BioFIND is a cross-sectional study of a cohort of which participants were enrolled at 8 sites in the United States.^6^ The average duration of PDs in this cohort is 8.34 years, which is much longer than the PD duration of subjects in the PPMI, and enrolled patients were receiving symptomatic treatment. Data downloaded from https://biofind.loni.usc.edu on April 25, 2018, were used for this analysis. A total of 126 moderate-advanced PD participants were included for analysis.

We incorporated a wide range of phenotypic features for analysis, including motor and non-motor manifestations based upon validated rating scales, biomarkers, and brain imaging data. In addition, other features including demographics, genetic risk score, and medications were also included. The details of all features used for analysis are listed in the appendix (supplemental Table 1, p 3-5), in which we have matched features collected between PPMI and BioFIND as much as possible. Specifically, we used sub-scores of the Movement Disorder Society Unified Parkinson Disease Rating Scale (MDS-UPDRS) Part I, extracting responses to individual questions regarding pain, fatigue, hallucinations, and apathy.^7^ We also analyzed composite scores for Tremor and Postural instability and gait disturbance (PIGD) as motor features.

**Table 1:** Experimental settings for generating phenotypic association graphs

| Setting No. | Dataset and sample selected | Features |
| --- | --- | --- |
| 1 | PPMI, PD baseline before symptomatic treatment, 1-5 years follow-up | Features valid in all periods of PPMI |
| 2 | BioFIND, PD baseline before symptomatic treatment, | Features valid in both BioFIND and PPMI baseline |
| 3 | PPMI, PD at baseline | Features valid in PPMI baseline |
| 4 | BioFIND, PD | Features valid in BioFIND |

PPMI=Parkinson's Progression Markers Initiative. BioFIND=Fox Investigation for New Discovery of Biomarkers. PD=Parkinson's disease.

### Phenotypic association graph generation

The PC algorithm was used to generate the association graphs of selected PD features.^8^ This algorithm starts from a fully connected graph, of which the nodes are all features studied and the undirected edges among them are candidate associations. Then it searches and determines which edges should be deleted by conditional independence testing. When the algorithm converges, we can obtain the phenotypic association graph of the clinical features. The detail of the algorithm is shown in the appendix (p 1).

In order to compare the associations among features in different cohorts (PPMI and BioFIND), as well as at different PD durations (within the PPMI cohort), we ran the PC algorithm upon different settings of dataset and features, as listed in Table 1. In particular, we first ran the PC algorithm on data from subjects at different times from baseline in PPMI to find the dynamic relationships among features valid in all periods of PPMI (Setting 1 in Table 1). For cross-cohort comparison, we ran the algorithm on BioFIND data with shared features of BioFIND and PPMI (Setting 2 in Table 1). Where there exist features valid only at baseline of PPMI and BioFIND, i.e., cerebrospinal fluid (CSF) biomarkers (including Aβ1-42, total tau [t-tau], phosphorylated tau [p-tau], and α-synuclein) and brain imaging data (including magnetic resonance imaging [MRI] and single photon mission computed tomography using ^131^I-ioflupane [DaTscan]), we ran the algorithm with Settings 3 and 4 in Table 1.

### Feature grouping based on graph structure

When generating phenotypic association graphs with longitudinal data from the PPMI study, there exists an inconsistent problem since the discussion of the feature-level would be too fine-grained to overcome noise. For example, two features may be linked directly at baseline and 2 years after baseline, while the edge may disappear at 1-year follow-up. This is due to the limited number of subjects of the studied cohorts. To address this issue, we partitioned the phenotypic features into different groups according to structure of the graphs and discuss the group-level relationships, which will be more consistent. Specifically, we first constructed a weighted summarization graph, of which nodes are all clinical features excluding demographics, medicine, biomarkers, and brain imaging features and the weight of each connection is the frequency of occurrence of the connection over all periods. Then the Louvain community detection algorithm,^9^ which aims at clustering nodes into densely intra-connected yet sparsely inter-connected groups, was utilized to group the features on the summarization graph (described in more detail in the appendix (p 2)).

## Results

### Phenotypic association graphs from PPMI

The phenotypic association graphs from PPMI were generated based on Setting 1 in Table 1. Figure 2 illustrates the generated graph at baseline. Seven feature groups were identified, including Motor, Mood, Cognitive-1, Cognitive-2, Autonomous dysfunction & sleepiness disorder (ADSD), Hallucinations, and Other daily symptoms (Table 2). We observed that clinical features in similar medical domains are grouped together. For example, MDS-T (tremor score), MDS-P (postural instability and gait disturbance score), MDS H&Y (Hoehn and Yahr) stage were grouped into the Motor, while GDS (Geriatric Depression Scale) and STAI (Geriatric Depression Scale) were grouped into the Mood. In addition, the fine-grained (i.e., feature-level) graphs at 1-5 years’ follow-up of PPMI are shown in the appendix (Supplemental Figures S1-5, p 6-8).

**Table 2:**
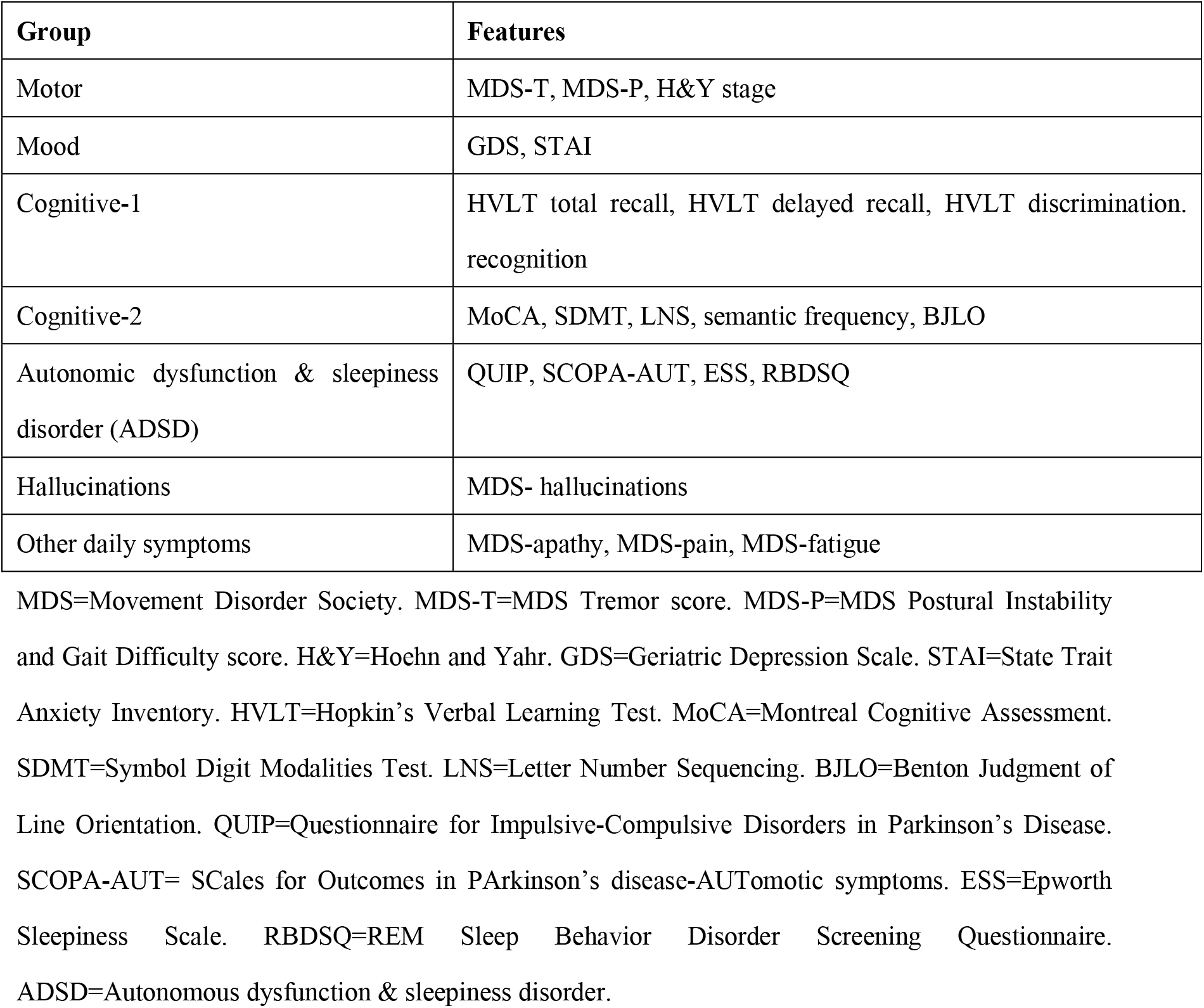
List of the feature groups obtained from the phenotypic association graph of PPMI cohort.

| Group | Features |
| --- | --- |
| Motor | MDS-T, MDS-P, H&Y stage |
| Mood | GDS, STAI |
| Cognitive-1 | HVLT total recall, HVLT delayed recall, HVLT discrimination. recognition |
| Cognitive-2 | MoCA, SDMT, LNS, semantic frequency, BJLO |
| Autonomic dysfunction & sleepiness disorder (ADSD) | QUIP, SCOPA-AUT, ESS, RBDSQ |
| Hallucinations | MDS- hallucinations |
| Other daily symptoms | MDS-apathy, MDS-pain, MDS-fatigue |

**Figure 2:**
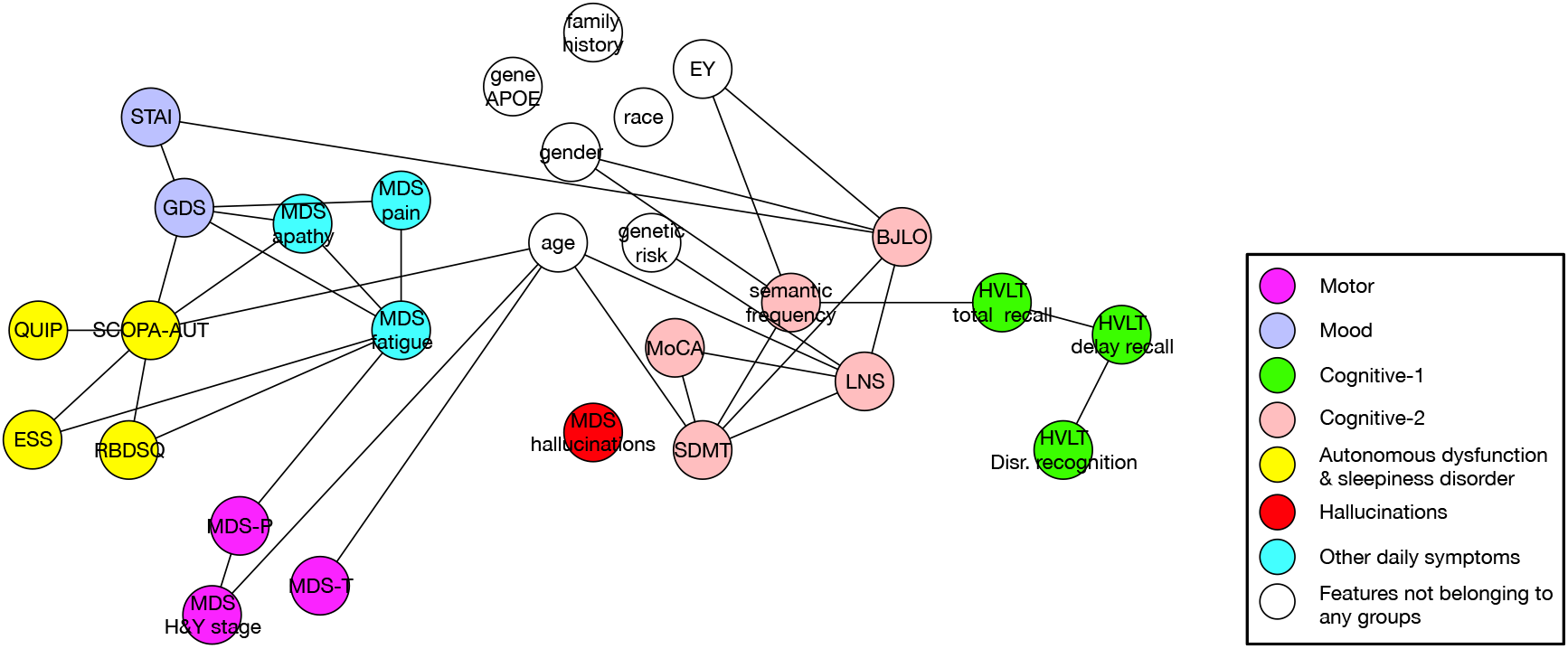
The feature-level phenotypic association graph at baseline of PPMI cohort. The selected samples and features follow setting 1 in Table 1. Each node represent a feature and colors of the nodes represent which group the specific feature belongs to. Nodes in white are those features which do not participate in grouping. MDS=Movement Disorder Society. MDS-T=MDS Tremor score. MDS-P=MDS Postural Instability and Gait Difficulty score. H&Y=Hoehn and Yahr. GDS=Geriatric Depression Scale. STAI=State Trait Anxiety Inventory. HVLT=Hopkin’s Verbal Learning Test. MoCA=Montreal Cognitive Assessment. SDMT=Symbol Digit Modalities Test. LNS=Letter Number Sequencing. BJLO=Benton Judgment of Line Orientation. QUIP=Questionnaire for Impulsive-Compulsive Disorders in Parkinson’s Disease. SCOPA-AUT=SCales for Outcomes in PArkinson’s disease-AUTomotic symptoms. ESS=Epworth Sleepiness Scale. RBDSQ=REM Sleep Behavior Disorder Screening Questionnaire. EY=Years of education.

By considering each feature group as a super-node, we generated the group-level association graphs of different periods in PPMI (see Figure 3). As shown in Figures 2, 3, and S1-5, connections between different groups are relatively sparse at baseline while more connections between groups emerge as PD progresses. To interpret these findings, we divide the edges between features into two types: (i) stable edges occurring throughout the different time periods examined, and (ii) longitudinal edges which change as the disease progresses (see Discussion section).

**Figure 3:**
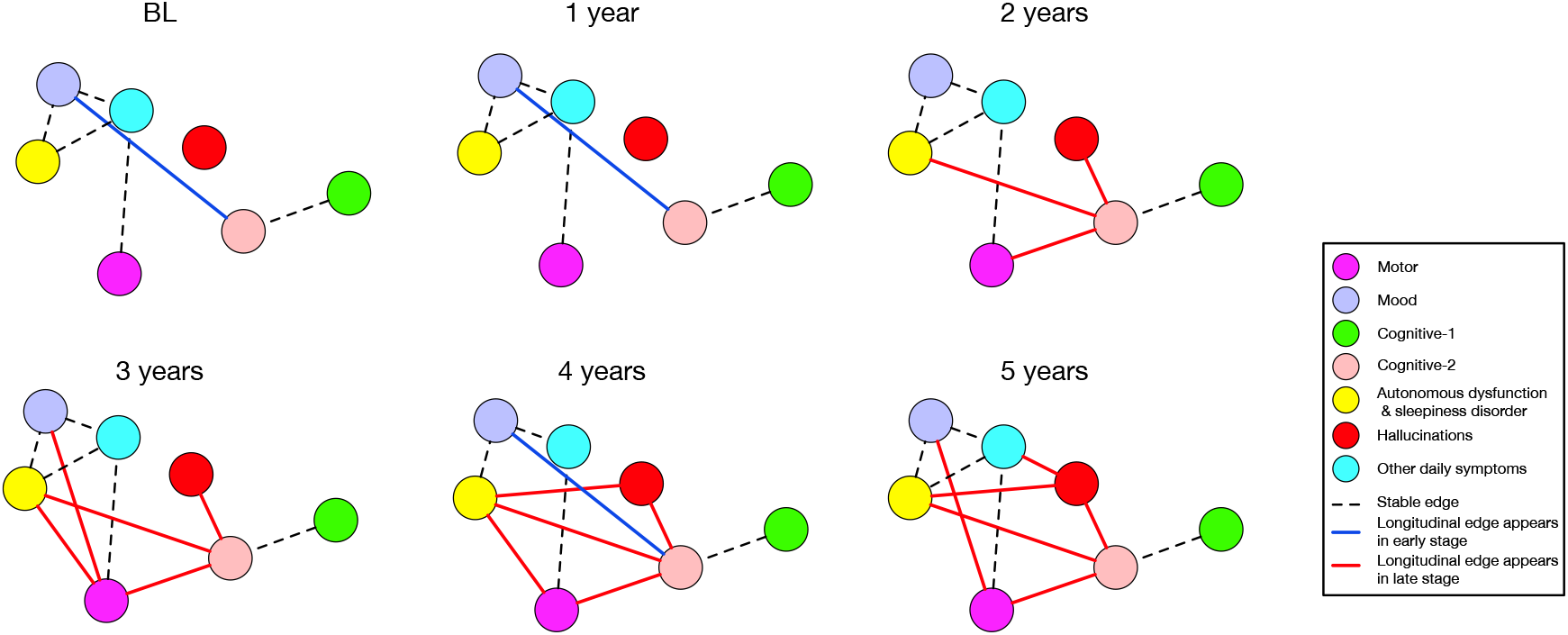
The group-level longitudinal phenotypic association graphs of PPMI cohort. The selected samples and features follow setting 1 in Table 1. Each node represent a extracted feature group. Stable edges (appear in all time points) are marked as dashed lines while longitudinal edges are marked in blue (appear in the early stage) or in red (appear in the late stage).

### Phenotypic association graph from BioFIND

Phenotypic association graph from BioFIND was generated based on Setting 2 in Table 1, shown in Figure 4. Six feature groups were identified, including Motor, Mood, Cognitive-2, Autonomous dysfunction & sleepiness disorder (ADSD), Hallucinations, and Other daily symptoms.

**Figure 4:**
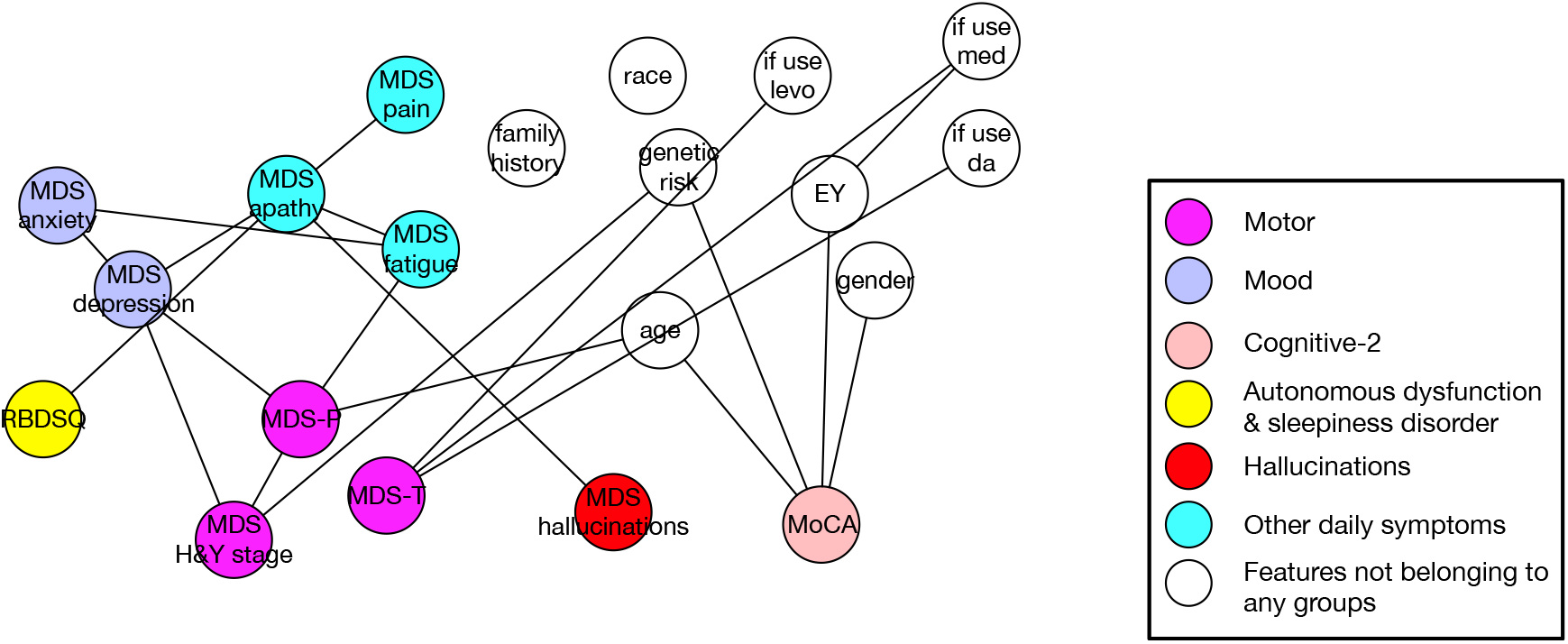
The feature-level phenotypic association graph of BioFIND study. Each node represent a feature and colors of the nodes represent which group the specific feature belongs to. Nodes in white are those features which do not participate in grouping. MDS=Movement Disorder Society. MDS-T=MDS Tremor score. MDS-P=MDS Postural Instability and Gait Difficulty score. H&Y=Hoehn and Yahr. GDS=Geriatric Depression Scale. MoCA=Montreal Cognitive Assessment. SDMT=Symbol Digit Modalities Test. LNS=Letter Number Sequencing. RBDSQ=REM Sleep Behavior Disorder Screening Questionnaire. EY=Years of education.

Except for some features of PPMI that are not available in the BioFIND study, the graph derived from BioFIND (see Figure 4) is similar to that generated from later-period data from PPMI (see Supplemental Figures S4 and 5). This is likely because patients within BioFIND have a longer duration of PD at enrollment, compared to the cohort enrolled in PPMI.

### Phenotypic association graphs with CSF biomarkers

To further investigate the correlations between CSF biomarkers and clinical features, we incorporated CSF biomarkers into our approach and generated the phenotypic association graphs of PPMI baseline data and BioFIND data according to Settings 3 and 4 in Table 1, respectively. For the PPMI cohort, we also incorporated features of MRI that provides structural information and DaTscan that provides information on integrity of the nigrostriatal tract. The resulting association graphs are shown in Figures 5 and 6.

**Figure 5:**
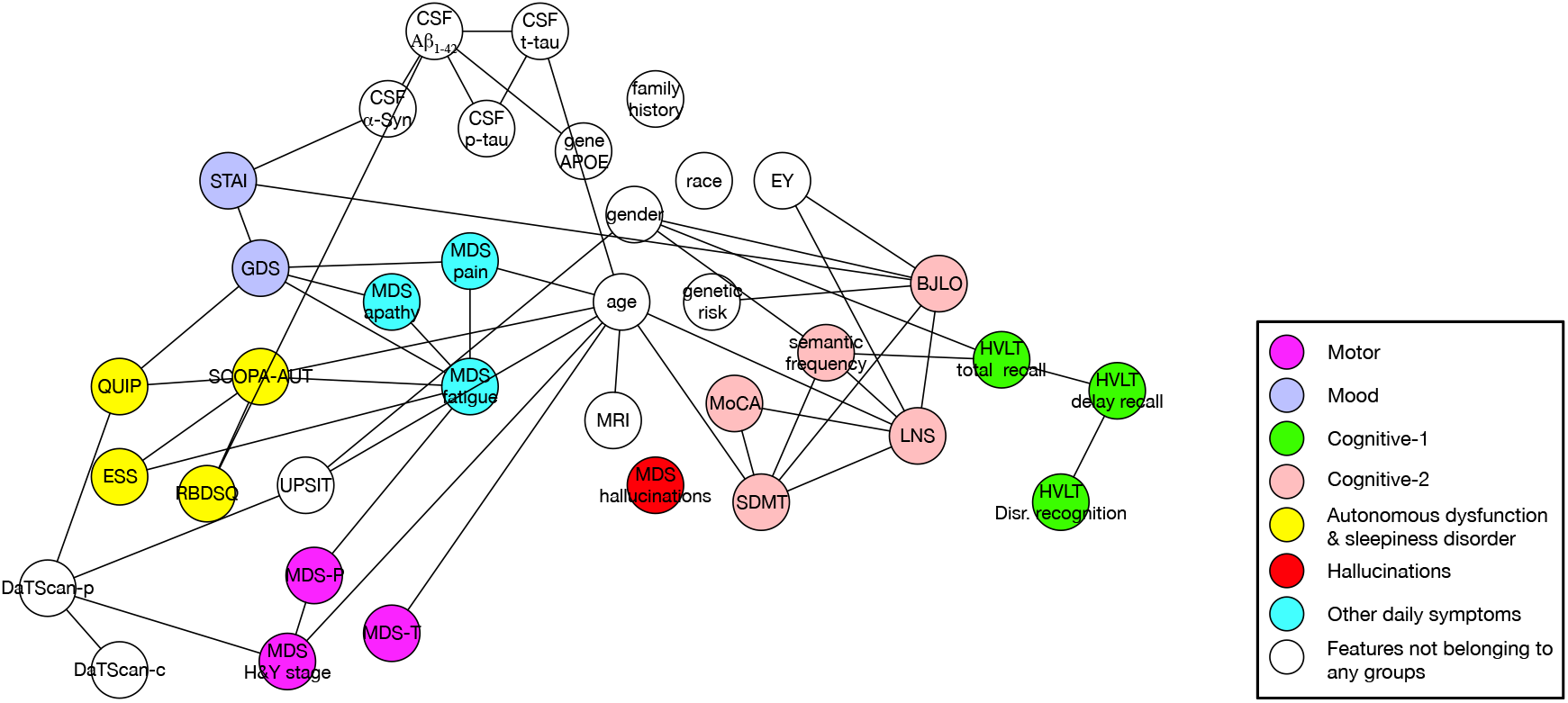
The feature-level phenotypic association graph at baseline of PPMI cohort, after incorporating CSF biomarkers and brain imaging. Each node represent a feature and colors of the nodes represent which group the specific feature belongs to. Nodes in white are those features which do not participate in grouping. MDS=Movement Disorder Society. MDS-T=MDS Tremor score. MDS-P=MDS Postural Instability and Gait Difficulty score. H&Y=Hoehn and Yahr. GDS=Geriatric Depression Scale. STAI=State Trait Anxiety Inventory. HVLT=Hopkin’s Verbal Learning Test. MoCA=Montreal Cognitive Assessment. SDMT=Symbol Digit Modalities Test. LNS=Letter Number Sequencing. BJLO=Benton Judgment of Line Orientation. QUIP=Questionnaire for Impulsive-Compulsive Disorders in Parkinson’s Disease. SCOPA-AUT=SCales for Outcomes in PArkinson’s disease-AUTomotic symptoms. ESS=Epworth Sleepiness Scale. RBDSQ=REM Sleep Behavior Disorder Screening Questionnaire. EY=Years of education. CSF= Cerebrospinal fluid. DaTscan-c=DaTscan Caudate. DaTscan-p=DaTscan Putamen.

**Figure 6:**
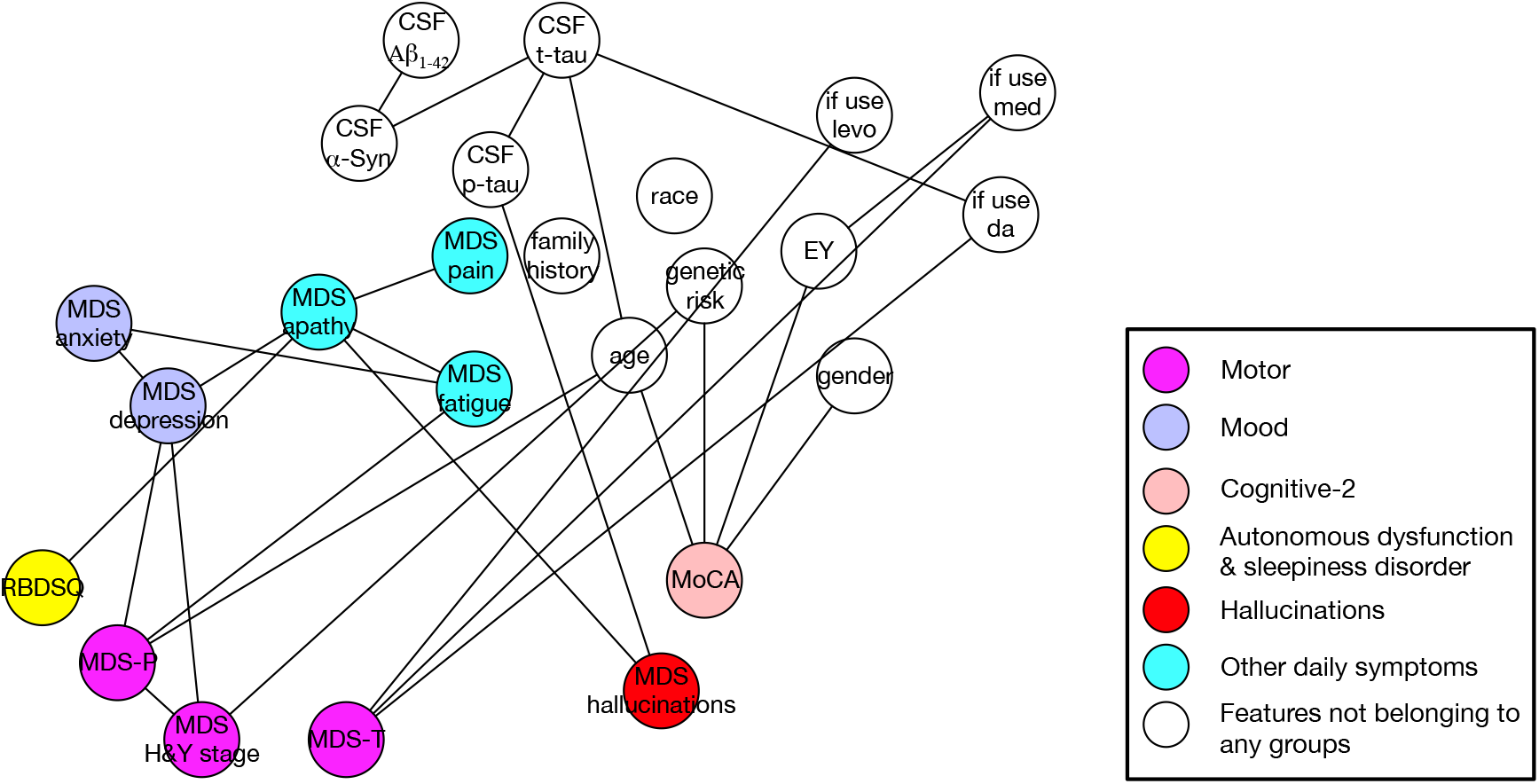
The feature-level phenotypic association graph of BioFIND after incorporating CSF biomarkers. Each node represent a feature and colors of the nodes represent which group the specific feature belongs to. Nodes in white are those features which do not participate in grouping. MDS=Movement Disorder Society. MDS-T=MDS Tremor score. MDS-P=MDS Postural Instability and Gait Difficulty score. H&Y=Hoehn and Yahr. GDS=Geriatric Depression Scale. MoCA=Montreal Cognitive Assessment. SDMT=Symbol Digit Modalities Test. LNS=Letter Number Sequencing. RBDSQ=REM Sleep Behavior Disorder Screening Questionnaire. EY=Years of education. CSF= Cerebrospinal fluid.

Generally, incorporating CSF biomarkers and brain imaging did not change the overall structure of the graph significantly. At PPMI baseline, CSF biomarkers are associated with mood (STAI) and sleepiness (RBDSQ) symptoms, age, and gene-APOE, while MRI is only associated with age. For BioFIND, the CSF biomarkers are associated with cognitive (MoCA) and hallucination symptoms. We will discuss more details in the Discussion section.

## Discussion

### Stable phenotypic associations in PD

From the graphs shown in Figures 2-6 and S1-5, we can conclude the stable phenotypic associations as follows:

1. We identified stable associations between Cognitive-1 and 2 groups. This is consistent with the features in the two groups all serving as measures of cognitive manifestations.
2. We identified stable associations between Mood and Other daily symptoms (including apathy, pain, and fatigue), and feature-level intra-group and inter-group connections such as the associations between GDS and STAI, between GDS and MDS-apathy, and between MDS-apathy and MDS-fatigue. The potential reasons could be two-fold: on one hand, measures of depression, anxiety, apathy, and fatigue have overlapping content in their respective scale questionnaires; on the other hand, depression, anxiety, apathy and fatigue likely share pathology in PD, being associated with the production of dopamine and serotonin and other neurochemicals.^10–12^
3. We identified stable associations between Mood and ADSD features. These two groups contain common non-motor neurodegenerative features of PD: depression (GDS), anxiety (STAI), REM sleep behavior disorder (RBDSQ), excessive daytime sleepiness (ESS), impulsive-compulsive disorders (QUIP), and autonomic dysfunction (SCOPA-AUT). They have a critical impact on the patient’s health-related quality of life. Besides the group-level connections, there exist consistent feature-level connections, such as associations between GDS and SCOPA-AUT, ESS and SCOPA-AUT, as well as RBD and SCOPA-AUT. Previous studies have reported that these features are correlated with the severity of PD and are mostly correlated with each other.^1–4^ Based upon PPMI data, any two of these features are associated with each other significantly (p-value < 0.01). The findings could potentially be explained by the spread of neurodegeneration within the brainstem. Braak et al reported that Lewy pathology and neuronal loss have been identified in the locus coeruleus, raphe nuclei, dorsal motor nucleus of the vagus (DMV) and pedunculopontine nuclei (PPN) simultaneously as PD processes.^13^ These regions are implicated in the control of daytime sleepiness, mood, and autonomic function.^2,3,14^ Other pathological studies also supported such associations.^15,16^ Our results add strength to the evidence that lesions in these regions can act as confounders of these dysfunctions. In addition, we also detected stable associations between ESS and RBDSQ, within the ADSD group, which could be explained by that they share close pathology.^17^ Moreover, SCOPA-AUT acts as a bridge connecting depression and sleepiness features: SCOPA-AUT is directly connected to GDS, RBD, ESS at all time points, while GDS is independent to RBDSQ or ESS and conditioned on SCOPA-AUT. One explanation would be that SCOPA-AUT, as the measurement of autonomic dysfunction, plays the role of a good proxy variable of the above neurodegeneration process, since it covers different aspects of autonomic functions (i.e., in both central nervous system and peripheral nervous system). Other studies have found a direct correlation between depression and sleep problems which may be attributable to that a) there exist a pathological relationship between the two sets of features, b) there are methodological issues based on the subject’s self-completed questionnaires (since patients with depression may tend to endorse numerous symptoms); c) the usage of antidepressant would cause RBD or other sleepiness problems.^18^ Moreover, the fact that both ESS and RBDSQ show conditional independence to GDS in our structure suggests that these effects may not be significant in the PPMI data.
4. The use of anti-PD medications is linked to features in ADSD group. Though feature-level connections may not be so consistent, we can see SCOPA-AUT, ESS and RBDSQ are connected to PD medicine. This could potentially be explained by the side effect profile of PD medications, as they may lead to orthostatic hypotension, excessive daytime drowsiness and sleep disruption.^19^

### Longitudinal phenotypic associations in PD

Base on the phenotypic association graphs generated at different periods within PPMI, we identified associations that vary over time:

1. An association between the features of the Motor and Cognitive-2 groups begins to occur at 2-5 years after baseline. In particular, at the feature-level, there is an edge connecting MDS-P and SDMT/MoCA at 3-5 years after baseline. This suggests that a common neurodegenerative process may be causing cognitive dysfunction (especially processing speed and attentional dysfunction measured by SDMT) and postural instability and gait disturbance (PIGD) phenotype. Such neurodegenerative processes may include degeneration of dopaminergic systems affecting parallel basal ganglia-thalamocortical pathways and non-dopaminergic degeneration within cholinergic systems.^20,21^
2. Associations connecting motor symptoms to ADSD and Mood symptoms emerge at two years after baseline. In particular, at the feature-level, there are connections between SCOPA-AUT and MDS-P, and between GDS and MDS-P. According to previous clinical studies, non-motor features comprising mood, sleep, and autonomic dysfunction are particularly related to PIGD phenotype. For example, PD patients of the PIGD subtype are more likely to have depression,^22^ REM sleep behavior disorder,^23^ and autonomic dysfunction than those of the tremor dominant subtype.^24^ In addition, the relation connecting GDS and MDS-P could be explained by a common pathology affecting the locus coeruleus, since this structure is implicated in the control of mood and in features of the PIGD subtype including postural stability.^2,25^ The edge between SCOPA-AUT and MDS-P is likely due to the degeneration within noradrenergic and cholinergic systems. As noted above, the PIGD phenotype is associated with cholinergic degeneration, and autonomic dysfunction in PD is also linked to cholinergic pathways which may include structures such as the DMV and PPN.^26^ Since there exist connections between SCOPA-AUT and MDS-P, and between GDS and MDS-P, there probably exist multiple non-dopaminergic pathologies which are related to noradrenergic and cholinergic systems. Although sleep problems (REM sleep behavior disorder and excessive daytime sleepiness) are also found to be correlated to the PIGD phenotype, RBDSQ or ESS are independent of MDS-P and conditioned on other variables such as SCOPA-AUT in our results. This suggests that there is likely no shared pathway between sleep problem and the PIGD phenotype. The correlation between them arises from parallel pathways, some of which affect autonomic function, depression, and sleep and some of which affect autonomic functions, depression, and PIGD phenotype.
3. We also discovered connections between the ADSD and Cognitive-2 groups in the later periods analyzed (2-5 years after baseline), especially between SCOPA-AUT and SDMT (Symbol Digit Modalities Test). The finding is consistent with existing clinical studies that there is no correlation between autonomic dysfunction and cognitive impairment at early-stage PD patients but there is a correlation between them with more advanced patients.^27,28^ Exact pathological correlations between autonomic dysfunction and cognitive decline have not been established, only one previous work explains this α-synuclein accumulation in the central and peripheral autonomic nervous systems as well as in neocortical areas.^28^ Other studies have focused on the relationship between cognitive function and orthostatic hypotension, a specific type of autonomic dysfunction.^29^ However, when we broke down SCOPA-AUT into different sub-scores (i.e., gastrointestinal, urinary, orthostatic, thermoregulatory, pupillomotor, and sexual), we found that gastrointestinal and urinary scores are also dependent on SDMT in advanced periods, even when conditioned on orthostatic scores and other features. This indicates that the correlation between autonomic dysfunction and cognitive impairment may derive from pathways associated with multiple aspects of autonomic dysfunction. For example, recurrent episodic hypotension may result in cerebral hypoperfusion, and in turn cause subcortical ischemic damage and subsequent cognitive impairment;^29^ and degeneration of the nervous system affects different aspects of autonomic functions. If we combine this with the above connections between SCOPA-AUT and MDS-P, and between MDS-P and SDMT/MoCA, we obtain triangle linkages among SCOPA-AUT, MDS-P, and cognitive features (i.e., MoCA and SDMT). Interestingly, such linkages start to be significant approximately during the same periods (beginning 2-3 years after baseline). These simultaneous relations may indicate shared pathologies among autonomic dysfunction, PIGD phenotype, and cognitive function (especially processing speed and attentional function), possibly due to degeneration within the cholinergic system as suggested by the above results. According to Braak’s staging analysis, Lewy pathology and neuronal loss in the DMV and pedunculopontine nuclei (PPN) begins in the early stages of PD.^13^ This contrasts with our correlations among autonomic function, PIGD and cognitive function, that are not found in the early-stage data in PPMI. Therefore, further research to address this contrast is needed.
4. There are relationships between Hallucinations and ADSD and Cognitive-2 groups in relatively late-stage patients, especially the feature-level relations between hallucinations and SCOPA-AUT, and between hallucinations and MoCA/SDMT/BJLO. As for the connection between hallucinations and cognitive impairment, multiple structural and functional neuroimaging studies have reported overlap in brain regions abnormalities in PD patients with hallucinations and those with cognitive impairment.^30^ The correlation between autonomic dysfunction and hallucinations was also reported in earlier studies.^31,32^ Although some previous studies explained it by more severe autonomic dysfunction reflecting more advanced PD disease,^31^ our result suggests that there potentially exist a more direct pathology affecting both autonomic dysfunction and hallucination, such as pathology of the DMV and other brainstem nuclei in addition to dysfunction of peripheral autonomic nerves.^32^ Moreover, the connection between hallucinations and autonomic dysfunction is likely to be at least in part a consequence of cholinergic degeneration discussed in the last three paragraphs.
5. The connection between Hallucinations and Apathy is observed at the last time point in this study (5 years after baseline). Though this correlation was reported in previous studies,^33^ there is no clear explanation yet and this deserves further investigation.
6. Although the genetic risk score is a good predictor of PD, there is no significant correlation between genetic risk score and motor features or non-motor symptoms. However, the genetic risk score is linked to cognitive features in the early stage of PD and this correlation disappears in more advanced periods (3-5 years after baseline). This could be explained by that: on one hand, the genetic risk score is associated with the cognitive functions;^34^ on the other hand, as the disease progresses, more other non-genetic factors affecting cognition arise, hence the genetic effect becomes “diluted” and not significant enough to be detected.
7. An inconsistent association exists between the Mood and Cognitive-2 groups at baseline, and 1 and 4 years after baseline, respectively. In the early stage, there exists feature-level connections between STAI and BJLO/LNS, while in the later stage the connection between GDS and MoCA emerges. Actually, the correlation between depression/anxiety and cognitive impairment has been frequently reported.^35^ One of the possible mechanisms underlying such a relationship is striatal dopamine degeneration.^36^ Another study reported the associations of cognitive impairment with both depression and cholinergic deficits and suggested that cholinergic deficits could underlie depression. This is supported by the finding that cortical cholinergic denervation was found to be associated with depression in patients with PD, independent of their cognitive functioning.^37^ This is reasonable according to our findings above. In particular, the connection between GDS and MoCA appears 4 years after baseline, coincident in time with the triangle connections among SCOPA-AUT, MDS-P, and MoCA/SDMT. The relation between anxiety and cognitive impairment may arise from different pathways since it occurs in the early stage of PD, which merits further investigation.

### Cross-cohort comparison between PPMI and BioFIND cohorts

As shown in Figures 4, S4, and S5, the phenotypic association graph in BioFIND is similar to those of PPMI at the later-timepoints. There are edges between age/gender/years of education and cognitive function (MoCA), between MDS-depression and MDS-anxiety, between MDS-depression and MDS-apathy, and between MDS-fatigue and MDS-apathy in the result of BioFIND. All of them are also presented as stable edges in PPMI. One inconsistent finding is that MDS-depression and RBDSQ are independent conditioned on MDS-apathy in BioFIND, while GDS and RBDSQ should be dependent even conditioned on MDS-apathy with SCOPA-AUT not included. If we replace GDS with MDS-depression in PPMI, we also found MDS-depression and RBDSQ are independent conditioned on MDS-apathy. This indicates that MDS-depression measured with a single question may not be reliable compared to GDS. In addition, there are connections between MDS-P and MDS-depression, MDS-apathy and MDS-hallucinations in BioFIND. These are consistent with the findings within PPMI. Our results are therefore consistent since BioFIND includes PD patients with more advanced disease.

### Phenotypic associations between CSF biomarkers and clinical features

As shown in Figures 5 and 6, we identified new associations after incorporating CSF-biomarker and brain imaging features for analysis. Besides strong connections among different CSF biomarkers, there are relations connecting CSF biomarkers with RBDSQ and STAI. The relation between anxiety and CSF biomarkers has also been reported in previous work on PPMI and an Alzheimer’s disease cohort.^38,39^ This suggests that there is potentially a direct effect of α-synuclein pathology on particular areas of the brain or that those neurochemical deficits (such as neurotransmitter deficits) are involved in anxiety.^39^ The connection between CSF-Aβ1-42 and RBDSQ may indicate a potential β-Amyloid pathway causing RBD, which has not been well-studied. Moreover, the path connecting gene-APOE, CSF-Aβ1-42 and RBDSQ suggests that APOE can be the genetic factor causing RBD problem.

We also observed a connection between DaTscan-p (DaTscan Putamen) and H&Y stage. This shows that DaTscan results can not only indicate the existence but also the severity of PD. DaTscan and smelling are usually combined to detect PD, and the connection between UPSIT and DaTscan-p suggests that dopamine transporter deficit and olfactory dysfunction are not independent factors of PD and there would be a complex pathway between them. The dopamine transporter deficit may cause olfactory dysfunction or they both act as surrogates for adverse influences on dopaminergic cells from some other source.^40^

In BioFIND, analogous to our findings in PPMI, connections within CSF biomarkers and between age and CSF t-tau were detected. Yet, no direct edge between Aβ1-42 and t-tau was identified. In addition, the relations between CSF biomarkers and other clinical features are different with that within PPMI. This may be due to different stages of PD. However, we cannot verify this with current data, because that CSF biomarker data for the advanced stage in PPMI are not available.

## Data Availability

The data that support the findings of this study are openly available in PPMI (http://www.ppmi-info.org) and BioFIND (https://biofind.loni.usc.edu)

## Acknowledgments

The research is supported by Michael J. Fox Foundation grant number 14858, 14858.01 and 15914. Part of the data used in the preparation of this article were obtained from the Parkinson’s Progression Markers Initiative (PPMI) database (http://www.ppmi-info.org/data). For up-to-date information on the study, visit http://www.ppmi-info.org. PPMI–a public-private partnership–is funded by the Michael J. Fox Foundation for Parkinson’s Research and funding partners, including Abbvie, Avid, Biogen, Bristol-Mayers Squibb, Covance, GE, Genentech, GlaxoSmithKline, Lilly, Lundbeck, Merk, Meso Scale Discovery, Pfizer, Piramal, Roche, Sanofi, Servier, TEVA, UCB and Golub Capital.

## Reference

1. Neikrug AB, Maglione JE, Liu L, et al. Effects of sleep disorders on the non-motor symptoms of Parkinson disease. J Clin Sleep Med 2013; 9: 1119–1129.

2. Simuni T, Caspell-Garcia C, Coffey C, et al. Correlates of excessive daytime sleepiness in de novo Parkinson’s disease: a case control study. Mov Disord 2015; 30: 1371–1381.

3. Cui SS D. JJ, Fu R, et al. Prevalence and risk factors for depression and anxiety in Chinese patients with Parkinson disease. BMC Geriatr 2017; 17: 270.

4. Arnao V, Cinturino A, Valentino F, et al. In patient’s with Parkinson disease, autonomic symptoms are frequent and associated with other non-motor symptoms. Clin Auton Res 2015; 25: 301–307.

5. Marek K, Jennings D, Lasch S, et al. The parkinson progression marker initiative (PPMI). Prog Neurobiol 2011; 95: 629–635.

6. Kang UJ, Goldman JG, Alcalay RN, et al. The BioFIND study: characteristics of a clinically typical Parkinson’s disease biomarker cohort. Mov Disord 2016; 31: 924–932.

7. Goetz CG, Tilley BC, Shaftman SR, et al. Movement Disorder Society-sponsored revision of the Unified Parkinson’s Disease Rating Scale (MDS-UPDRS): scale presentation and clinimetric testing results. Mov Disord 2008; 23: 2129–2170.

8. Spirtes P, Glymour CN, Scheines R, et al. Causation, prediction, and search. Cambridge, Mass. : MIT Press, 2000.

9. Blondel VD, Guillaume JL, Lambiotte R, Lefebvre E. Fast unfolding of communities in large networks. J Stat Mech 2008; 2008: P10008.

10. Skorvanek M, Gdovinova Z, Rosenberger J, et al. The associations between fatigue, apathy, and depression in Parkinson’s disease. Acta Neurol Scand 2015; 131: 80–87.

11. Pavese N, Metta V, Bose SK, Chaudhuri KR, Brooks DJ. Fatigue in Parkinson’s disease is linked to striatal and limbic serotonergic dysfunction. Brain 2010; 133: 3434–3443.

12. Yamanishi T, Tachibana H, Oguru M, et al. Anxiety and depression in patients with Parkinson’s disease. Intern Med 2013; 52: 539–545.

13. Braak H, Ghebremedhin E, Rüb U, Bratzke H, Del Tredici K. Stages in the development of Parkinson’s disease-related pathology. Cell Tissue Res 2004; 318: 121–134.

14. Benarroch EE, Schmeichel AM, Sandroni P, Low PA, Parisi JE. Involvement of vagal autonomic nuclei in multiple system atrophy and Lewy body disease. Neurology 2006; 66: 378–383.

15. Schulz-Schaeffer WJ. The synaptic pathology of α-synuclein aggregation in dementia with Lewy bodies, Parkinson’s disease and Parkinson’s disease dementia. Acta Neuropathol 2010; 120: 131–143.

16. Boeve BF, Silber MH, Saper CB, et al. Pathophysiology of REM sleep behaviour disorder and relevance to neurodegenerative disease. Brain 2007; 130: 2770–2788.

17. Zhou J, Zhang J, Lam SP, et al. Excessive daytime sleepiness predicts neurodegeneration in idiopathic REM sleep behavior disorder. Sleep 2017; 40: zsx041.

18. Mahlknecht P, Seppi K, Frauscher B, et al. Probable RBD and association with neurodegenerative disease markers: a population-based study. Mov Disord 2015; 30: 1417–1421.

19. Chung S, Bohnen NI, Albin RL, Frey KA, Müller ML, Chervin RD. Insomnia and sleepiness in Parkinson disease: associations with symptoms and comorbidities. J Clin Sleep Med 2013; 9: 1131–1137.

20. Kelly VE, Johnson CO, McGough EL, et al. Association of cognitive domains with postural instability/gait disturbance in Parkinson’s disease. Parkinsonism Relat Disord 2015; 21: 692–697.

21. Bohnen NI, Albin RL. The cholinergic system and Parkinson disease. Behav Brain Res 2011; 221: 564–573.

22. Dissanayaka NN, Sellbach A, Silburn PA, O’Sullivan JD, Marsh R, Mellick GD. Factors associated with depression in Parkinson’s disease. J Affect Disord 2011; 132: 82–88.

23. Chahine LM, Amara AW, Videnovic A. A systematic review of the literature on disorders of sleep and wakefulness in Parkinson’s disease from 2005 to 2015. Sleep Med Rev 2017; 35: 33–50.

24. Alves G, Larsen JP, Emre M, Wentzel-Larsen T, Aarsland D. Changes in motor subtype and risk for incident dementia in Parkinson’s disease. Mov Disord 2006; 21: 1123–1130.

25. Grimbergen YA, Langston JW, Roos RA, Bloem BR. Postural instability in Parkinson’s disease: the adrenergic hypothesis and the locus coeruleus. Expert Rev Neurother 2009; 9: 279–290.

26. Sulzer D, Surmeier DJ. Neuronal vulnerability, pathogenesis, and Parkinson’s disease. Mov Disord 2013; 28: 715–724.

27. Idiaquez J, Benarroch EE, Rosales H, Milla P, Ríos L. Autonomic and cognitive dysfunction in Parkinson’s disease. Clin Auton Res 2007; 17: 93–98.

28. Malek N, Lawton MA, Grosset KA, et al. Autonomic dysfunction in early Parkinson’s disease: results from the United Kingdom tracking Parkinson’s study. Mov Disord Clin Pract 2017; 4: 509–516.

29. McDonald C, Newton JL, Burn DJ. Orthostatic hypotension and cognitive impairment in Parkinson’s disease: causation or association?. Mov Disord 2016; 31: 937–946.

30. Melzer TR, Watts R, MacAskill MR, et al. Grey matter atrophy in cognitively impaired Parkinson’s disease. J Neurol Neurosurg Psychiatry 2012; 83: 188–194.

31. Zhu K, van Hilten JJ, Putter H, Marinus J. Risk factors for hallucinations in Parkinson’s disease: results from a large prospective cohort study. Mov Disord 2013; 28: 755–762.

32. Barrett MJ, Smolkin ME, Flanigan JL, Shah BB, Harrison MB, Sperling SA. Characteristics, correlates, and assessment of psychosis in Parkinson disease without dementia. Parkinsonism Relat Disord 2017; 43: 56–60.

33. Santangelo G, Trojano L, Vitale C, et al. A neuropsychological longitudinal study in Parkinson’s patients with and without hallucinations. Mov Disord 2007; 22: 2418–2425.

34. Mata IF, Leverenz JB, Weintraub D, et al. APOE, MAPT, and SNCA genes and cognitive performance in Parkinson disease. JAMA Neurol 2014; 71: 1405–1412.

35. Alzahrani H, Venneri A. Cognitive and neuroanatomical correlates of neuropsychiatric symptoms in Parkinson’s disease: A systematic review. J Neurol Sci 2015; 356: 32–44.

36. Weintraub D, Newberg AB, Cary MS, et al. Striatal dopamine transporter imaging correlates with anxiety and depression symptoms in Parkinson’s disease. J Nucl Med 2005; 46: 227–232.

37. Bohnen NI, Kaufer DI, Hendrickson R, Constantine GM, Mathis CA, Moore RY. Cortical cholinergic denervation is associated with depressive symptoms in Parkinson’s disease and parkinsonian dementia. J Neurol Neurosurg Psychiatry 2007; 78: 641–643.

38. Kang JH, Mollenhauer B, Coffey CS, et al. CSF biomarkers associated with disease heterogeneity in early Parkinson’s disease: the Parkinson’s Progression Markers Initiative study. Acta Neuropathol 2016; 131: 935–949.

39. Ramakers I, Verhey FRJ, Scheltens P, et al. Anxiety is related to Alzheimer cerebrospinal fluid markers in subjects with mild cognitive impairment. Psychol Med 2013; 43: 911–920.

40. Doty RL. Olfaction in Parkinson’s disease and related disorders. Neurobiol Dis 2012; 46: 527–552.

